# Cardiovascular Adverse Events After Definitive Chemoradiotherapy for Lung Cancer in an Appalachian Population: Incidence and Machine Learning-Based Prediction

**DOI:** 10.64898/2026.04.01.26349944

**Authors:** Vivian Salama, Joseph A. Schmidlen, J. Christopher Knoth, Troy Nguyen, A. Naveen Joseph, Matthew Trotta, Ramon Alfredo Siochi, Raymond R. Raylman, Jeffrey Ryckman, Mohammed Almubarak, David A. Clump, Christopher M. Bianco, Mina F. Hanna, Phillip M. Pifer

## Abstract

**Background:** Cardiovascular adverse events (CVAEs) after chemoradiotherapy (CRT) for lung cancer are major concerns in Appalachia due to high rates of smoking and pre-existing cardiovascular diseases (CVD). The objectives of this study were to characterize the incidence of CVAEs in this population and evaluate machine learning (ML) models for CVAEs risk stratification and mortality prediction.

**Methods:** A retrospective study was conducted among Appalachian patients with lung cancer treated with definitive CRT at a single institution between 2013 and 2025. Baseline clinical variables, including demographics, smoking status, pre-existing CVD, and post-CRT CVAEs were collected. Heart dosimetric parameters were also obtained. ML-models [Random Forest (RF), Gradient Boosting (GBM), Support Vector Machine (SVM), Logistic Regression (LR)] were trained using 5-fold cross validation and evaluated using AUC, sensitivity, specificity, and F1 score. Feature importance was assessed using permutation analysis. Wilcoxon and Chi-squared tests were used for descriptive comparisons.

**Results:** Eighty-six patients (mean age 66 years, 47% male) were included. At diagnosis, 80% (n=69) had NSCLC and 20% (n=17) had LS-SCLC. CVAEs occurred in 51 patients (59%). The most frequent events were NSTEMI (n=15, 29.4%), pericardial disease (n=15, 29.4%), and arrhythmia (n=8, 15.7%). Mean heart dose was higher in the CVAE group (13.4 vs 9.4 Gy, *p=0.27*).

For CVAE prediction, GBM achieved the highest AUC (0.55, 95% CI 0.44-0.69) and sensitivity (75%), while RF showed the highest sensitivity (80%, 95% CI 69-90%). Key predictors included age and cardiac dosimetrists (Heart V20, V40, V50, and mean heart dose).

For mortality prediction, RF achieved the highest discrimination (AUC = 0.63, 95% CI 0.496-0.750). Age, cardiac dosimetry, disease stage, and cardiovascular comorbidity were the most influential predictors.

**Conclusion:** High incidence of CVAEs occurred among patients with lung cancer treated with CRT in this Appalachian cohort. While ML models demonstrated modest predictive performance, tree-based approaches demonstrated high sensitivity for identifying patients at risk for CVAEs and mortality. Age and cardiac radiation dose metrics consistently emerged as key predictors, highlighting the importance of cardiac dose optimization and ML-based risk stratification for cardio-oncology surveillance.

## 1 Introduction

Lung cancer remains the leading cause of cancer-related mortality in the United States (^1^). Radiotherapy (RT) is commonly utilized in the curative-intent setting. Definitive chemoradiotherapy (CRT) serves as a cornerstone of treatment and has demonstrated improved overall survival (OS) in patients with locally advanced non-small cell lung carcinoma (LA-NSCLC) (^2^) and limited-stage small cell lung carcinoma (LS-SCLC) (^3^). Because standard-of-care treatment has improved outcomes, and many risk factors for lung cancer and cardiovascular (CV) events overlap, understanding post-treatment toxicity becomes increasingly important. Due to proximity of normal organs to the primary tumor site, patients receiving CRT are at risk of developing side effects during and after treatment, including esophageal, pulmonary, and cardiovascular toxicity (CVT) (^4^).

Radiation-induced CVT is a well-recognized and clinically important consequence of RT in thoracic malignancies (^5–7^). Treatment-related CVT can manifest acutely, or months to years after exposure. CVTs include but are not limited to ischemic events, arrhythmia, congestive heart failure (CHF), and valvular disease (^8^). Studies investigating cardiac toxicity were primarily conducted in breast, esophageal, and Hodgkin lymphoma survivors (^9–11^). In lung cancer RT, exposure of the heart to ionizing radiation is almost inevitable to provide adequate tumor coverage. Moreover, patients with lung cancer often carry a significant smoking history and pre-treatment CV comorbidity that has been associated with CV complications following treatment (^12^). The multifactorial nature of radiation-induced CVT in lung cancer presents a challenging topic in treatment planning and risk stratification.

Patient factors and dosimetric relationships to CVT in patients receiving CRT have been investigated (^12, 13^); however, variability among studies regarding baseline cardiovascular disease (CVD) may underestimate risk in populations with high pre-treatment CVD burden. In a combined analysis of four prospective trials for LA-NSCLC, pre-existing cardiac disease was associated with a nearly three-fold risk of developing grade >3 cardiac events (^12^). However, only a quarter of patients in this study had pre-existing cardiac disease, limiting applicability to populations with higher CV risk. Another study found that a higher mean heart dose (MHD) correlated with worse OS; however, specific cardiac event endpoints were not investigated (^14^). Both cohorts were primarily treated with contemporary 3D-conformal radiotherapy (3D-CRT), which may overestimate risk when compared to more modern techniques, such as intensity modulated radiation therapy (IMRT).

Uncertainty in the current literature may be particularly relevant in Appalachia, a region that carries high rates of CVD (^15, 16^) tobacco smoking (^17^), and lung cancer incidence (^1, 17^). Patients in this region often carry a substantial baseline CV risk profile, raising concerns that existing literature may underestimate the true burden of CVT in underserved populations. Traditional regression-based approaches, which assume linear relationships and prespecified interactions, may not fully capture the complex interplay among clinical comorbidities, radiation dosimetry, and survival outcomes. Artificial Intelligence and machine learning (AI/ML) approaches offer the potential to model nonlinear relationships and higher-order interactions, thereby enhancing risk stratification. Emerging evidence supports the application of ML techniques in predicting radiation-associated CV outcomes (^18, 19^), yet data specific to Appalachian populations are lacking.

Post-CRT cardiovascular adverse events (CVAE) may arise from multiple contributing factors, including baseline CVD, systemic therapy, and potential RT-related cardiac exposure. To our knowledge, the current literature has not yet characterized post-CRT-CVAEs in an Appalachian lung cancer patient cohort. Improved risk stratification in our population can assist our clinicians in developing more individualized treatment plans, provide clearer perspective for clinicians practicing in other underserved regions, and present an opportunity for risk reduction within cardio-oncology. Therefore, the objectives of this study were to characterize the incidence of post-CRT-CVAEs and evaluate ML models for post-RT CV risk stratification and survival prediction in Appalachian patients with lung cancer treated with CRT. In this study, prediction models were developed using baseline clinical characteristics and treatment-related variables available at the time of CRT planning to estimate the risk of CVAEs during post-treatment follow-up. We hypothesized that this patient population would have a high burden of CVAEs after CRT and that incorporating clinical and dosimetric factors into ML-models could help identify high-risk patients early.

## 2 Materials and Methods

### 2.1 Study Design and Data Source

This institutional review board (IRB)–approved retrospective cohort study included adult patients with biopsy-confirmed lung cancer treated with definitive CRT at our institution between 2013 and 2025.

Eligible patients were identified through an institutional radiation oncology electronic medical record systems (i.e. ARIA and Eclipse) query. Clinical and demographic data were extracted from the integrated electronic health record (i.e., Epic) and treatment planning systems.

This study was reported in accordance with the Transparent Reporting of a multivariable prediction model for Individual Prognosis Or Diagnosis plus Artificial Intelligence (TRIPOD+AI) reporting guidelines for development and validation of prediction models (^20, 21^).

### 2.2 Study Population

Patients were included if they met the following **inclusion criteria**: (1) were ≥18 years of age, (2) had biopsy-proven lung cancer, (3) were treated with definitive-intent CRT to the primary lung tumor and mediastinal nodal metastasis if present, (4) had complete treatment and dosimetric data available, (5) were treated using contemporary RT techniques, including 3D-CRT, IMRT, or volumetric modulated arc therapy (VMAT). Eligible histology included: LA-NSCLC, LS-SCLC and select oligometastatic NSCLC treated with definitive CRT.

Patients were excluded if they met any of these exclusion criteria: (1) received non-definitive or palliative treatment, (2) had lung metastases from a non-pulmonary primary tumor, (3) had early stage NSCLC treated with surgery, stereotactic body radiotherapy (SBRT), or hypofractionated RT, (4) received more than one prior course of thoracic RT before a documented CVAE (to avoid confounding from cumulative dose exposure). Patients were followed from the final day of RT until first documented CV event, death **or** last known clinical follow-up. The median follow-up was 30.5 months (IQR 13.75-47). Cardiovascular events occurring during active radiation treatment were assigned a time-to-event of 0 months.

After applying eligibility criteria, 86 patients were included in the final analysis. A study flow diagram is presented in Figure 1. The study sample included all eligible patients treated during the study period and therefore no formal sample size calculation was performed.

**Figure 1:**
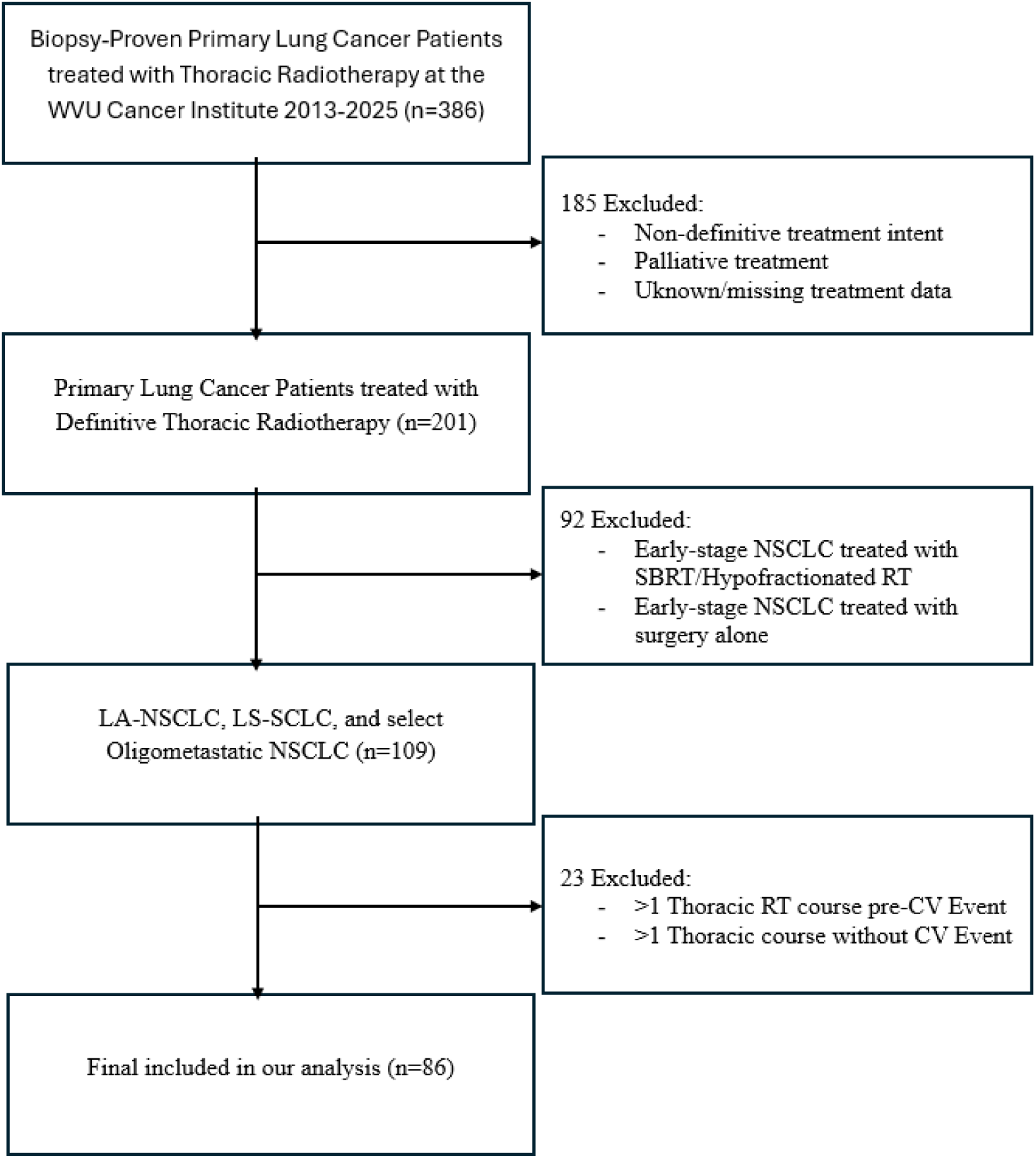
Flow Chart of Study Population.

### 2.3 Predictors/Variables

Candidate predictors were prespecified based on clinical relevance and prior literature. Predictors included: age at diagnosis, sex, and race/ethnicity, smoking status (current, former, never), pre-existing CV comorbidity (defined as having the presence of any coronary artery disease, heart failure, hypertension, cerebrovascular accident, peripheral artery disease, or arrhythmia before receiving RT), primary tumor features (site, histology and staging), treatment variables (e.g., surgery, chemotherapy, immunotherapy, total RT dose and number of fractions) and cardiac dosimetric parameters extracted from the treatment planning system [Mean heart dose (MHD), heart V20(%), heart V30(%), heart V40 (%), and heart V50 (%)]. Whole heart is auto contoured in the CT simulation planning DICOM files and whole heart dosimetrics parameters extracted from Eclipse. Figure 2 represents the finalized radiation treatment plan conducted in our institution. All predictor variables were measured prior to or at the time of CRT planning.

**Figure 2:**
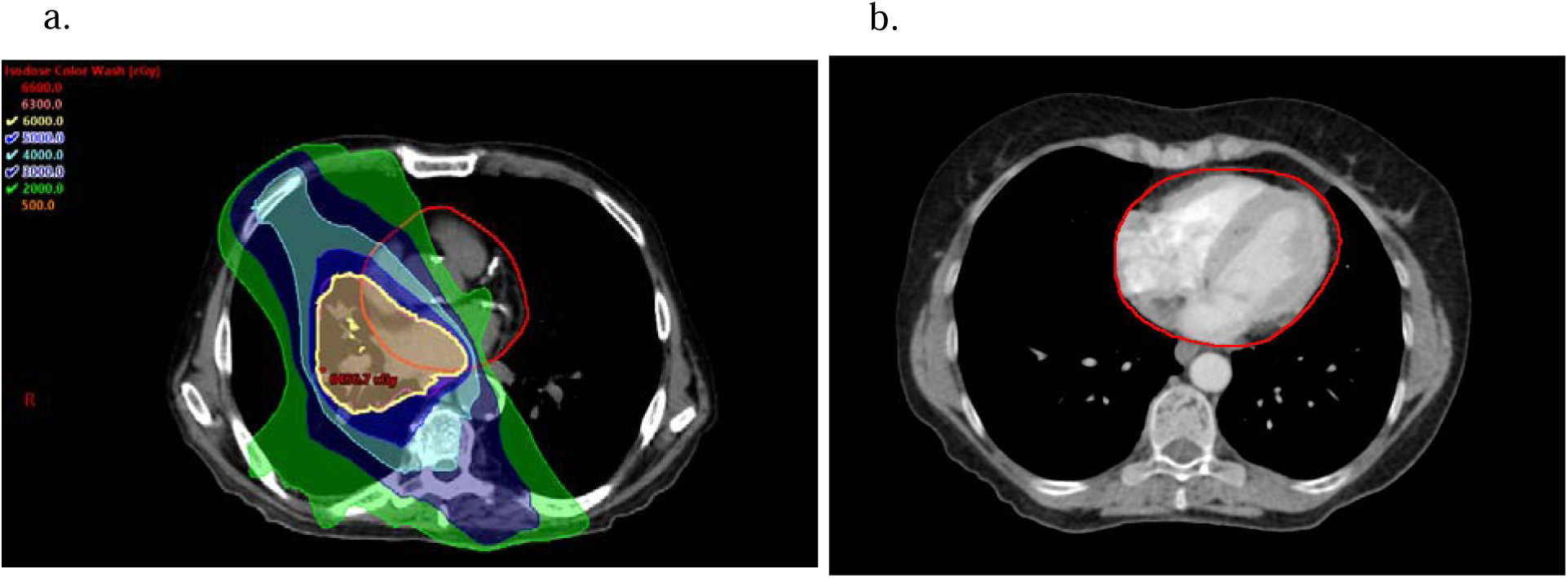
External beam radiation treatment plan in lung cancer. a. Non contrast CT simulation planning of an Appalachian patient diagnosed with lung cancer (NSCLC) showing the 60 (yellow), 50 (dark blue), 40 (light blue), 30 (purple), 20 (green) Gy Isodose Lines (IDLs) w/ PTV (pink) and Whole heart contour (red). b. Contrast CT image of another patient showing whole heart contours (red).

### 2.4 Outcome Definition

The primary outcome in this study was the development of any CVAE during or after radiation treatment, dichotomized into [0 = No-CVAE and 1 = Yes-CVAE]. Cardiac events that were recorded included, non-ST-elevation myocardial infarction (NSTEMI), ST-elevation myocardial infarction (STEMI), arrhythmia, CHF, pericardial disease, cardiac death, new cardiology referral, valvular dysfunction, new clinical diagnosis of CAD, or other. Events in the “other” category could include any major structural or physiologic change to the heart (e.g. ventricular aneurysm, reduced ejection fraction). Events were confirmed if patients received a formal diagnosis in the EHR and by documented physician diagnoses or imaging reports.

The secondary outcome was patient mortality, dichotomized as (0 = alive and 1 = deceased). Survival was determined using institutional EHRs and confirmed through documented date of death, when applicable. Survival time was calculated from the completion of RT to death or last known follow-up.

### 2.5 Statistical Analysis

#### 2.5.1 Descriptive statistics

Descriptive analysis of patient characteristics was performed by calculating the number (n), percent (%) for the categorical features and the mean and standard deviation (SD) or median and Inter-Quantile Range (IQR) of the numeric features, as appropriate. Differences in patient characteristics between CVAE and mortality were compared using Wilcoxon test for numeric variables and the Chi-square test or Fisher Exact Test (if a cell count is <5) for categorical variables. A 2-sided *P-value* of less than 0.05 was considered statistically significant.

#### 2.5.2 Machine Learning models

##### 2.5.2.1 Preprocessing

Twenty variables were primarily used for ML model training. Categorical variables included: sex, race/ethnicity, smoking, CV comorbidity, tumor histology, site of primary tumor, overall stage, T stage, N Stage, M Stage, immunotherapy and surgery. Numerical variables included: age at diagnosis, RT dose, RT fractions, MHD, and heart V20, V30, V40, and V50.

Missing values were calculated and variables with ≥ 10% missing data were excluded. Separate preprocessing pipelines were constructed for numerical and categorical features. For numerical variables, missing values were imputed using the median of each variable, and variables were then standardized using z-score normalization via StandardScaler. For categorical variables, missing values were imputed using the most frequent value in each column, then variables were ordinally encoded using Ordinal Encoder, with unknown categories during inference encoded as -1. Preprocessing was implemented using a column transformer and embedded within a scikit-learn to prevent data leakage.

Class imbalance was evaluated prior to model training. When imbalance between outcome classes was identified, the Synthetic Minority Over-sampling Technique (SMOTE) (^22, 23^) was applied to mitigate class imbalance.

Preprocessing steps were embedded within the modeling pipeline to ensure that all transformations were performed separately within each training fold.

##### 2.5.2.2 Classification model training and validation

Four ML classification models were investigated in Python including tree-based Random Forest (RF), tree-based Gradient Boosting Machine (GBM), Logistic Regression (LR), and Support Vector Machine (SVM). All models were initialized with default settings with hyper-parameter optimization. Logistic regression models were regularized using L2 penalty; the regularization strength (*C = 0.01*) controlled coefficient shrinkage to reduce overfitting, with the “lbfgs” solver and a maximum of iterations (*max_iter = 2000*), and class weights = “balanced.”. Random forest models were constructed using an ensemble of decision trees, with hyperparameters including number of trees (*n_estimators = 200*), maximum tree depth (*max_depth = None*), and minimum samples per leaf (*min_samples_leaf =4*), minimum samples per split (*min_samples_split = 2*) and class weight = None. Gradient boosting models sequentially built shallow trees, with learning rate (*learning_rate = 0.1*), number of estimators (*n_estimators = 100*), and maximum tree depth (*max_depth = 2*) and subsampling (*subsample = 1.0*). The SVM model utilized a radial basis function (RBF) kernel, with hyperparameters *C = 0.1* and gamma = “scale,” controlling margin regularization and kernel curvature, respectively.

Hyperparameters were either tuned using cross-validated grid search set to default values, and model complexity was constrained to mitigate overfitting given the small sample size.

Model development and internal validation were performed using 5-fold stratified cross-validation with shuffling (random seed=42). For each candidate classifier, out-of-fold predicted probabilities for the CVAE outcome were generated using cross_val_predict, ensuring that each patient’s risk estimate was produced by a model not trained on that patient.

##### 2.5.2.3 Model Evaluation

Models’ performance evaluation was conducted using discrimination metrics and calibration. Discrimination was assessed using the area under the receiver operating characteristic curve (AUC), sensitivity, specificity, and F1-score. Uncertainty was quantified using nonparametric bootstrapping (2000 resamples) applied to out-of-fold predictions to generate 95% confidence intervals for all performance metrics.

Calibration was assessed using out-of-fold predicted probabilities obtained from 5-fold stratified cross-validation. Calibration curves were generated by grouping predictions into eight quantile-based bins and comparing the mean predicted probability within each bin to the observed event rate. Perfect calibration was represented by the 45-degree reference line. Calibration plots were generated for each model using internally validated predictions. External validation was not performed due to limited sample size and will be addressed in future multi-institutional studies.

##### 2.5.2.4 Model explainability and Feature importance

Model explainability assessed using permutation feature importance for the best-performing tree-based classifiers. Following preprocessing, each model was refitted on the full dataset to obtain a final fitted estimator for interpretability analyses. Permutation importance was computed by randomly permuting each predictor and quantifying the resulting decrease in model discrimination, measured by the AUC. Each permutation was repeated 30 times, and the mean and standard deviation of the AUC decrease were reported. Features with larger mean AUC decreases were interpreted as contributing more strongly to model performance. Permutation feature importance was visualized for the top 15 predictors in each model using horizontal bar plots. Importance values represent the mean decrease in AUC following random permutation of each feature, with error bars corresponding to the standard deviation across 30 permutation repetitions. Features were ranked by mean AUC decrease.

Scikit-learn packages were used for ML modeling, validation, and evaluation. All statistical analyses were performed using Python 3.12, JMP PRO 15 and Graph Pad Prism.

## 3 Results

### 3.1 Descriptive analysis

A total of 86 patients with lung cancer treated with definitive CRT at our institution were included in the study. The cohort included 46 females (53%) and 40 males (47%). The mean age at diagnosis was 66 years (SD = 9.13; 95% CI: 64.0–68.0), with a median age of 67.5 years (IQR: 59–72) and a range of 39 to 87 years. The median RT total dose was 60 Gy (range: 44–70 Gy; IQR: 60–60), with a mean dose of 58.7 Gy (SD = 6.12; 95% CI: 57.4–60.0). The median number of fractions was 30 (range: 22–35; IQR: 30–30), with a mean of 30 fractions (SD = 1.78; 95% CI: 29.82–30.59). Systemic therapy regimens included 49% (n=42) patients received carboplatin/paclitaxel, 13% (n=11) received cisplatin/etoposide, 12% (n=10) received carboplatin/etoposide, 9% (n=8) received carboplatin/pemetrexed, 6% (n=5) received carboplatin only, 2% (n=2) received carboplatin/etoposide/pembrolizumab, 1% (n=1) received carboplatin/pemetrexed/pembrolizumab, 2% (n=2) did not receive systemic therapy, and 6% (n=5) received multiagent systemic therapy. During any point in treatment, 77% (n=66) received immunotherapy.

Overall, 51 patients (59%) developed CVAE during or after RT, whereas 35 patients (41%) did not experience a CVAE. At the time of last follow-up, 34 patients (40%) were alive, and 52 patients (60%) were deceased.

Among the 51 patients who developed CVAE during or after RT, the most common events were NSTEMI in 15 patients (29.41%) and pericarditis or pericardial effusion in 15 patients (29.41%), followed by arrhythmia in 8 patients (15.69%). Less frequent events included CHF in 4 patients (7.84%), and valvular dysfunction in 2 patients (3.92%). Other CVAEs included mild reduction in ejection fraction (1 patient, 1.96%), cardiomyopathy (1 patient, 1.96%), and left ventricular apical aneurysm (1 patient, 1.96%) (Figure 3).

**Figure 3:**
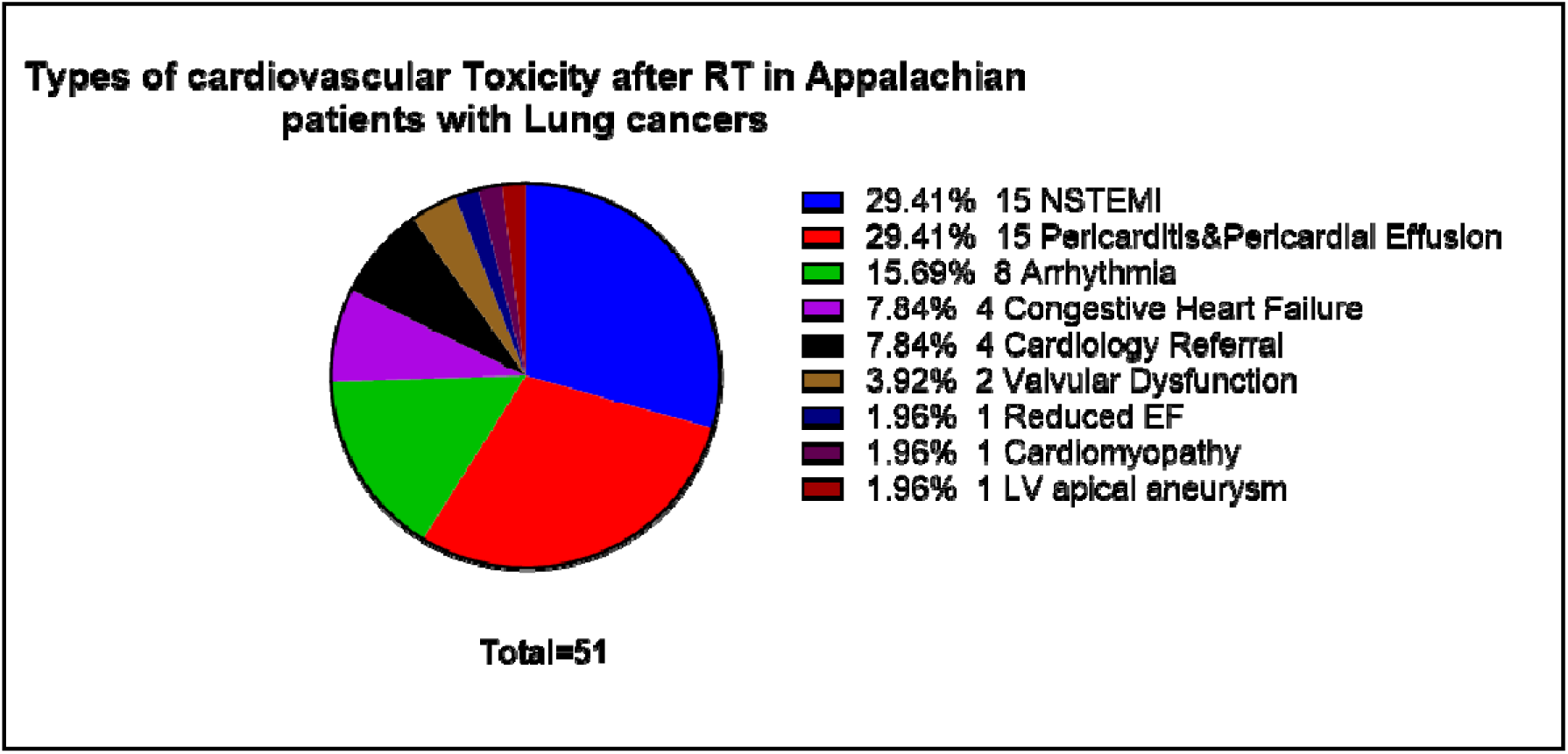
Frequency and types of cardiovascular adverse events during and after RT in Appalachian patients with lung cancer.

Cardiac dose volume parameters showed: mean heart dose (Gy), Median=(11.8), Range=0.06-32.24, IQR=5.70-14.88; Heart V50 (%): Median=3.72, Range=0-33.33, IQR=0.03-10.73; Heart V40 (%): Median=3.72, Range=0-33.33, IQR=1.03-10.73; Heart V30 (%): Median=9.77, Range=0-50.07, IQR=2.36-17.51 and Heart V20 (%): Median=20.31, Range=0-67.24, IQR=6.255-29.89.

Table 1 provides a summary of the cohort characteristics and the results of the Chi-square and Wilcoxon tests.

**Table 1:** Descriptive and characteristic table of patients with lung cancers treated with RT, according to cardiovascular events during or after RT and survival/mortality status.

| Variables | All Patients (n = 86) | No CVT (n = 35) | Yes CVT (n = 51) | *P value* | Alive (n = 34) 40% | Deceased (n = 52) 60% | *P value* |
| --- | --- | --- | --- | --- | --- | --- | --- |
| **Age at Diagnosis median, (IQR)**<br>**Age mean, (SD)** | 67.5, (59-72)<br>66, SD=9.13 | 66, (59-74)<br>66.5, (SD=10.05) | 68, (60-72)<br>65.6, (SD=8.5) | 0.901 | 60.5, (56-70)<br>62.6, (SD=8.8) | 69, (61-73)<br>68.25, (SD=8.7) | 0.004* |
| **Sex, n (%)** |  |  |  | 0.751 |  |  | 0.934 |
| Male | 40 (47%) | 17 (20%) | 23 (27%) |  | 16 (19%) | 24 (28%) |  |
| Female | 46 (53%) | 18 (21%) | 28 (33%) |  | 18 (21%) | 28 (32%) |  |
| **Race, n (%)** |  |  |  | 0.564 |  |  | 0.821 |
| White 0 | 83 (97%) | 33 (39%) | 50 (58%) |  | 33 (38%) | 50 (59%) |  |
| Black 1 | 3 (3%) | 3 (2%) | 1 (1%) |  | 1 (1%) | 2 (2%) |  |
| **Smoking** |  |  |  | 0.427 |  |  | 0.821 |
| Never | 4 (5%) | 1 (1%) | 3 (3%) |  | 1 (1%) | 3 (3%) |  |
| Former | 57 (66%) | 21 (24%) | 36 (42%) |  | 23 (27%) | 34 (39%) |  |
| Current | 25 (29%) | 13 (15%) | 12 (14%) |  | 10 (12%) | 15 (17%) |  |
| **Cardiovascular comorbidity** |  |  |  | 0.630 |  |  | 0.008* |
| No | 22 (25%) | 8 (9%) | 14 (16%) |  | 14 (16%) | 8 (9%) |  |
| Yes | 64 (74%) | 27 (31%) | 37 (43%) |  | 20 (23%) | 44 (51%) |  |
| **Primary cancer type/Histology, n (%)** |  |  |  | 0.621 |  |  | 0.499 |
| Adenocarcinoma | 25 (29%) | 11 (13%) | 14 (16%) |  | 11 (13%) | 14 (16%) |  |
| Squamous Cell Carcinoma | 39 (45%) | 16 (18%) | 23 (27%) |  | 14 (16%) | 25 (29%) |  |
| Small Cell Neuroendocrine Carcinoma | 17 (20%) | 5 (6%) | 12 (14%) |  | 8 (9%) | 9 (10%) |  |
| Large Cell Neuroendocrine Carcinoma | 1 (1%) | 1 (1%) | 0 (0%) |  | 0 (0%) | 1 (1%) |  |
| Not otherwise specified | 1 (1%) | 1 (1%) | 0 (0%) |  | 1 (1%) | 0 (0%) |  |
| Poorly Differentiated Carcinoma | 3 (3%) | 1 (1%) | 2 (2%) |  | 0 (0%) | 3 (3%) |  |
| **Site of Primary Tumor, n (%)** |  |  |  | 0.221 |  |  | 0.423 |
| Right lung | 43 (50%) | 21 (24%) | 22 (26%) |  | 18 (21%) | 25 (29%) |  |
| Left lung | 42 (49%) | 14 (16%) | 28 (33%) |  | 15 (17%) | 27 (31%) |  |
| Mediastinal LN with unknown origin | 1 (1%) | 0 (0%) | 1 (1%) |  | 0 (0%) | 1 (1%) |  |
| **Stage at Diagnosis, n (%)** |  |  |  | 0.7629 |  |  | 0.5575 |
| Stage I | 1 (1%) | 0 (0%) | 1 (1%) |  | 1 (1%) | 0 (0%) |  |
| Stage II | 5 (6%) | 1 (1%) | 4 (5%) |  | 2 (2%) | 3 (3%) |  |
| Stage III | 57 (66%) | 26 (30%) | 31 (36%) |  | 22 (25%) | 35 (41%) |  |
| Stage IV | 6 (7%) | 2 (2%) | 4 (5%) |  | 1 (1%) | 5 (6%) |  |
| Limited Stage (For Neuroendocrine Carcinomas) | 17 (20%) | 6 (7%) | 11 (13%) |  | 8 (9%) | 9 (10%) |  |
| **T stage, n (%)** |  |  |  | 0.856 |  |  | 0.343 |
| T1 | 20 (23%) | 9 (10%) | 11 (13%) |  | 8 (9%) | 12 (14%) |  |
| T2 | 16 (19%) | 6 (7%) | 10 (12%) |  | 9 (11%) | 7 (8%) |  |
| T3 | 19 (22%) | 6 (12%) | 13 (13%) |  | 5 (6%) | 14 (16%) |  |
| T4 | 21 (24%) | 10 (11%) | 11 (13%) |  | 7 (8%) | 14 (16%) |  |
| Tx | 4 (5%) | 1 (2%) | 3 (3%) |  | 3 (3%) | 1 (1%) |  |
| Not available–NA | 6 (7%) | 3 (3.5%) | 3 (3.5%) |  | 2 (2%) | 4 (5%) |  |
| **N stage, n (%)** |  |  |  | 0.398 |  |  | 0.613 |
| N0 | 9 (10%) | 3 (3%) | 6 (7%) |  | 2 (2%) | 7 (8%) |  |
| N1 | 9 (10%) | 5 (6%) | 4 (4%) |  | 2 (2%) | 7 (8%) |  |
| N2 | 33 (38%) | 15 (17%) | 18 (21%) |  | 15 (17%) | 18 (21%) |  |
| N3 | 25 (29%) | 7 (8%) | 18 (21%) |  | 12 (14%) | 13 (15%) |  |
| Nx | 4 (5%) | 3 (3%) | 1 (1%) |  | 1 (1%) | 3 (4%) |  |
| NA | 6 (7%) | 2 (2%) | 4 (5%) |  | 2 (2%) | 4 (5%) |  |
| **M Stage, n (%)** |  |  |  | 0.752 |  |  | 0.752 |
| M0/Mx | 74 (86%) | 30 (35%) | 44 (51%) |  | 31 (36%) | 43 (50%) |  |
| M1a | 0 (0%) | 0 (0%) | 0 (0%) |  | 0 (0%) | 0 (0%) |  |
| M1b | 4 (5%) | 2 (2.5%) | 2 (2.5%) |  | 1 (1%) | 3 (4%) |  |
| M1c | 2 (2%) | 0 (%) | 2 (2%) |  | 0 (%) | 2 (2%) |  |
| Not available–NA | 6 (7%) | 3 (3.5%) | 3 (3.5%) |  | 2 (2%) | 4 (5%) |  |
| **Status, n (%)** |  |  |  | 0.602 |  |  |  |
| Deceased-0 | 52 (61%) | 20 (23%) | 32 (37%) |  | Not Applicable | Not Applicable |  |
| Alive-1 | 34 (39%) | 15 (17%) | 19 (22%) |  | Not Applicable | Not Applicable |  |
| **CVAE, n (%)** |  |  |  |  |  |  | 0.602 |
| No CVAE | 35 (41%) | Not Applicable | Not Applicable |  | 15 (17%) | 20 (23%) |  |
| Yes CVAE | 51 (59%) | Not Applicable | Not Applicable |  | 19 (22%) | 32 (37%) |  |
| **Immunotherapy, n (%)** |  |  |  | 0.942 |  |  | 0.570 |
| No | 20 (23%) | 8 (9%) | 12 (14%) |  | 9 (10%) | 11 (13%) |  |
| Yes | 66 (77%) | 27 (32%) | 39 (45%) |  | 25 (29%) | 41 (48%) |  |
| **Surgery, n (%)** |  |  |  | 0.572 |  |  | 0.428 |
| No | 74 (86%) | 31 (36%) | 43 (50%) |  | 28 (33%) | 46 (53%) |  |
| Yes | 12 (14%) | 4 (5%) | 8 (9%) |  | 6 (7%) | 6 (7%) |  |
| **Mean Heart (Gy)** | Median=11.8, IQR=5.70-14.88 | Median=9.44, IQR=6.34-14.69 | Median=13.38, IQR=5.56-15.48 | 0.272<br>Median Test<br>(0.0495*) | Median=10.23, IQR=7.51-14.36 | Median=12.98, IQR=4.71-16.12 | 0.937 |
| **Heart V50 (%)** | Median=3.72, IQR=0.03-10.73 | Median=1.36, IQR=0.24-5.51 | Median=1.4, IQR=0-5.95 | 0.671 | Median=1.32, IQR=0.18-5.19 | Median=1.41, IQR=0.02-5.88 | 0.919 |
| **Heart V40 (%)** | Median=3.72, IQR=1.03-10.73 | Median=3.69, IQR=1.13-9.96 | Median=4.04, IQR=0.90-12.56 | 0.754 | Median=2.73, IQR=1.01-10.51 | Median=4.06, IQR=1.04-10.83 | 0.708 |
| **Heart V30 (%)** | Median=9.77, IQR=2.36-17.51 | Median=6.92, IQR=2.53-15.81 | Median=11.6, IQR=3.21-33.13 | 0.339 | Median=9.05, IQR=2.53-16.30 | Median=10.63, IQR=3.29-18.56 | 0.560 |
| **Heart V20 (%)** | Median=20.31, IQR=6.255-29.89 | Median=13.13, IQR=4.54-27.11 | Median=22.85, IQR=6.56-31.04 | 0.159<br>Median Test<br>(0.0203*) | Median=15.86, IQR=6.04-28.41 | Median=21.80, IQR=7.37-30.18 | 0.478 |

### 3.2 Machine Learning models performance

#### Machine Learning models for CVAE prediction and risk stratification during and after CRT in Appalachian lung cancer population

Table 2 summarizes the performance discriminative metrics among the ML models for CVAE prediction and risk stratification. Among the evaluated ML models, the GBM classifier demonstrated the highest discriminative performance, with an AUC of 0.567 (95% CI: 0.440–0.686). This model achieved a sensitivity of 0.73 (95% CI: 0.60–0.85), specificity of 0.29 (95% CI: 0.14–0.44), and an F1 score of 0.65 (95% CI: 0.54–0.75). The RF model achieved a close AUC of 0.514 (95% CI: 0.381–0.639) however, demonstrated the highest sensitivity among the evaluated classifiers (0.80; 95% CI: 0.69–0.91).

**Table 2:** Discrimination metrics and the performance of the models predicting CVAE during and after CRT in Appalachian lung cancer population.

| Model | AUC, 95% CI | Sensitivity, 95% CI | Specificity, 95% CI | F1, 95% CI |
| --- | --- | --- | --- | --- |
| Gradient Boosting Machine (GBM) | 0.567, [0.440, 0.686] | 0.725, [0.596, 0.846] | 0.286, [0.139, 0.438] | 0.655, [0.542, 0.752] |
| Logistic Regression (LR) | 0.524, [0.397, 0.646] | 0.588, [0.444, 0.727] | 0.457, [0.294, 0.622] | 0.600, [0.469, 0.706] |
| Random Forest Model (RF) | 0.514, [0.381, 0.639] | 0.804, [0.689, 0.909] | 0.171, [0.056, 0.297] | 0.678, [0.579, 0.768] |
| Support Vector Machine (SVM) | 0.492, [0.360, 0.622] | 0.725, [0.600, 0.840] | 0.314, [0.167, 0.485] | 0.661, [0.552, 0.759] |

The LR model showed modest discrimination with an AUC of 0.524 (95% CI: 0.397–0.646), with sensitivity of 0.59 (95% CI: 0.44–0.73) and specificity of 0.46 (95% CI: 0.29–0.62). The SVM (RBF kernel) model showed the lowest discrimination, with an AUC of 0.492 (95% CI: 0.360–0.622).

Calibration curvesdemonstrated variability in probability calibration across the evaluated models (Figure 4.a). The LR model showed the best calibration, indicating good agreement between predicted and observed event probabilities. In contrast, the RF and GBM demonstrated poorer calibration, with larger deviations from the ideal line.

**Figure 4:**
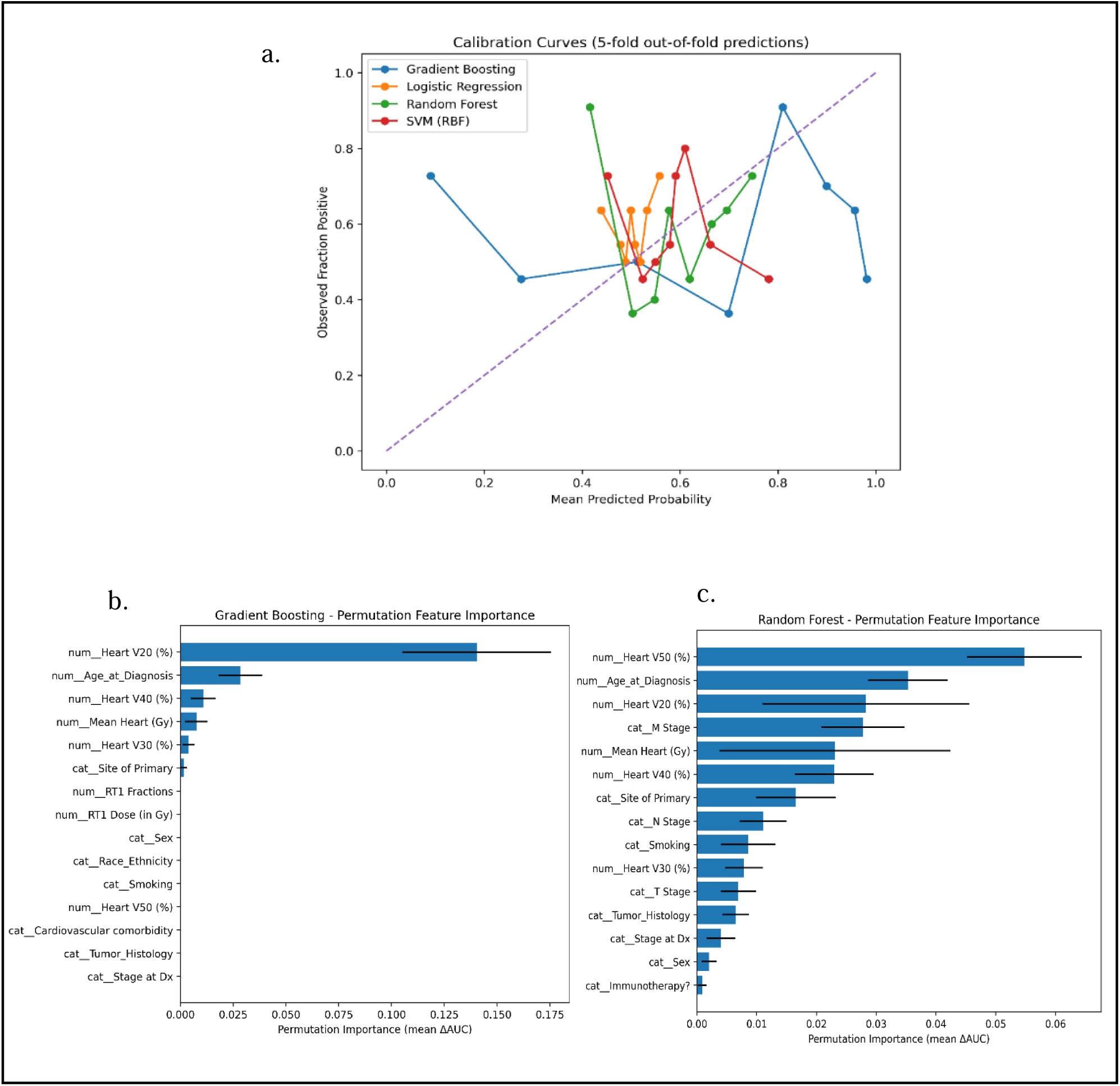
Calibration and feature importance of ML models predicting CVAEs during and after definitive CRT in lung cancer population a. calibration curve of ML models predicting CVAE. b. Feature importance of the top 15 features of Gradient Boosting Model using permutation feature importance. c. Feature importance of the top 15 features of Random Forest Model using permutation feature importance.

Feature importance of tree-based models (GBM and RF) was summarized in Figures 4.b, 4.c and Supplementary File S1. For GBM (Figure 4.b), heart V20 (%) emerged as the most important predictor, with the highest mean importance score (0.140 ± 0.035), indicating that this dosimetric parameter had the largest impact on model performance. Age at diagnosis was the second most influential feature (0.028 ± 0.010). Additional cardiac dosimetric variables, including heart V40 (%) (0.011 ± 0.006), MHD (0.008 ± 0.005), and heart V30 (%) (0.004 ± 0.003), showed smaller but measurable contributions to the model. Among categorical variables, site of primary tumor demonstrated minimal importance (0.002 ± 0.001).

In the RFM (Figure 4.c, Supplementary File S1), heart V50 (%) was identified as the most influential feature (0.055 ± 0.010), followed by age at diagnosis (0.035 ± 0.007) and heart V20 (%) (0.028 ± 0.017). Disease stage also contributed to model predictions, particularly M stage (0.028 ± 0.007) and N stage (0.011 ± 0.004). Additional dosimetric features, including MHD (0.023 ± 0.019) and heart V40 (%) (0.023 ± 0.007), demonstrated moderate importance. Other variables such as smoking status, site of primary tumor, and tumor histology showed small contributions.

##### Feature Selection

We evaluated the four ML models using a reduced set of the highly clinically and dosimetrically relevant and important variables (age at diagnosis, smoking status, pre-existing CV comorbidity, and cardiac dose metrics).

Discrimination remained limited across all models. The highest AUC was observed with GBM as well (AUC = 0.54), followed by RF (AUC = 0.52) and SVM (AUC = 0.51). Logistic Regression demonstrated slightly worse-than-chance discrimination (AUC = 0.48). Across all models, sensitivity was consistently higher than specificity. GBM achieved the most balanced performance (sensitivity = 0.76, specificity = 0.37), while RF and SVM showed higher sensitivity (>0.80) but low specificity (≤0.23). LR also demonstrated low specificity (0.20). F1 scores ranged from 0.64 (LR) to 0.70 (GBM), reflecting moderate classification performance driven largely by sensitivity rather than true class discrimination.

Importantly, the reduced-feature models achieved performance comparable to earlier models that included a larger number of variables, suggesting that the additional baseline features did not contribute meaningful predictive information.

#### Machine Learning models for mortality prediction and risk stratification after definitive CRT in Appalachian lung cancer population

Four ML models (RF, LR, GBM, and SVM) were utilized to predict mortality status (alive vs. deceased at last follow-up) using baseline clinical, tumor, treatment, and cardiac dosimetric variables.

Table 3 illustrates the mortality models’ performance metrics. Discriminative performance improved compared with prior CVAE models. RF achieved the highest discrimination (AUC = 0.63, 95% CI 0.496, 0.750), followed closely by LR (AUC = 0.61) (Figure 5.a). Sensitivity was consistently higher than specificity across all models. RF showed high sensitivity (0.90) but limited specificity (0.24). LR demonstrated the most balanced performance (sensitivity = 0.87, specificity = 0.38) and achieved the highest F1 score (0.76). Calibration curve is illustrated in Figure 5.b. Tree-based and linear models demonstrated modest but meaningful ability to distinguish mortality status using baseline variables.

**Table 3:** Discrimination metrics and the performance of the models predicting survival status after CRT in Appalachian lung cancer population.

| Model | AUC, 95% CI | Sensitivity, 95% CI | Specificity, 95% CI | F1, 95% CI |
| --- | --- | --- | --- | --- |
| Random Forest Model (RF) | 0.625, [0.496, 0.750] | 0.904, [0.816, 0.980] | 0.235, [0.102, 0.379] | 0.752, [0.656, 0.833] |
| Logistic Regression (LR) | 0.611, [0.480, 0.738] | 0.865, [0.761, 0.947] | 0.382, [0.214, 0.552] | 0.763, [0.667, 0.841] |
| Gradient Boosting Machine (GBM) | 0.529, [0.397, 0.657] | 0.673, [0.542, 0.800] | 0.441, [0.276, 0.611] | 0.660, [0.547, 0.759] |
| Support Vector Machine (SVM) | 0.404, [0.277, 0.532] | 0.981, [0.935, 1.000] | 0, [0.000, 0.000] | 0.744, [0.656, 0.822] |

**Figure 5:**
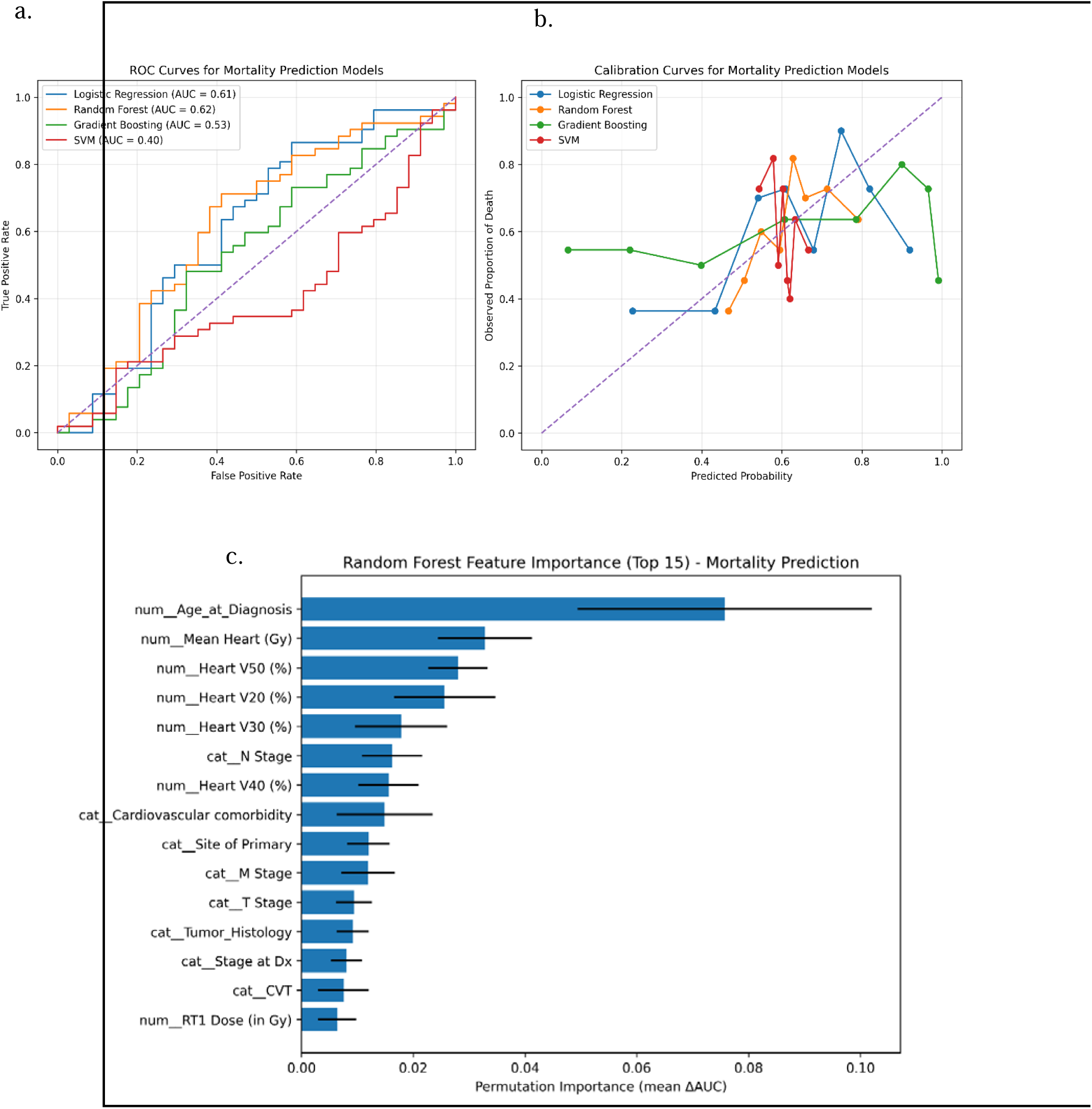
Performance and feature importance of ML models predicting mortality in patients with lung cancer treated with definitive CRT. a. Area under the Receiver Operating Characteristic Curve (AUC) of four ML models. b. Calibration curves of the four predictive models. c. feature importance of top 15 features of RF model predicting mortality in lung cancer population.

##### Feature Importance for Mortality Prediction

Permutation-based feature importance analysis was performed for the RF model as the highest performing model, to identify variables contributing most to mortality prediction (Figure 5.c). Age at diagnosis emerged as the most influential predictor (mean importance=0.076, SD=0.026). Among treatment-related factors, MHD (mean importance=0.033, SD=0.008) and cardiac dose volume metrics, particularly heart V50 (%), heart V20 (%), and heart V30 (%) were among the top contributors (mean importance= 0.028, 0.026, and 0.0179 respectively).

Disease-related staging variables also contributed meaningfully. N stage (mean=0.016, SD=0.005), M stage (mean=0.012, SD=0.005), and T stage (mean=0.009, SD=0.004) appeared among the more important categorical predictors. Baseline CV comorbidity was among the top clinical predictors (mean=0.015, SD=0.009). Tumor characteristics such as histology and site of primary tumor contributed to smaller but non-negligible predictive value. Interestingly, CVAE occurrence showed modest importance (mean=0.008, SD=0.005), suggesting that the development of CVT may be associated with increased mortality.

## 4 Discussion

In this retrospective study, post-CRT CVAEs were common in this Appalachian lung cancer cohort. To our knowledge, this is the first real-world, retrospective study investigating the incidence of CVAE and evaluating ML models for predicting CVAE and mortality using baseline clinical, therapeutic and cardiac dosimetric variables in this unique cohort of Appalachian lung cancer population received CRT. Our data analysis demonstrated important findings, including a high incidence of CVAEs (59%) during or after treatment, reflecting the high pre-treatment CVD burden in this population. Machine learning models demonstrated only modest discrimination for CVAE prediction but performed better for mortality prediction, with tree-based ensemble models demonstrating higher performance than linear regression models while regression models had more stable and balanced performance. Furthermore, cardiac radiation doses parameters and age at diagnosis consistently emerged as the most influential predictors across tree-based models, suggesting that radiation exposure to the heart and baseline patient characteristics play key roles in both CVAEs and survival outcomes.

The high incidence of CVAEs observed in our cohort isreflecting the unique characteristics of the Appalachian population. Appalachia is known to have disproportionately high rates of CVD, tobacco use, and socioeconomic health disparities, all of which contribute to increased baseline CV risk (^15–17^). Patients with lung cancer frequently have significant smoking histories and pre-existing CV comorbidities, which may amplify susceptibility to radiation-induced cardiac injury (^12^). The most common CVAEs observed in our study were NSTEMI and pericardial disease (pericarditis or pericardial effusion), followed by arrhythmia. These findings are consistent with prior studies demonstrating that thoracic radiation may lead to a spectrum of cardiac complications including ischemic heart disease, pericardial injury, and electrical conduction abnormalities (^24–26^). Dess et al., demonstrated that the incidence of grade ≥ 3 cardiac events exceeded 10% among patients with NSCLC treated with RT, additionally baseline cardiac disease and higher MHD were significantly associated with higher cardiac event rates (^12^). While Niedzielski et al. identified about 50% of patients with stage III NSCLC treated with RT developed grade 2 pericardial effusion. Importantly, although modern radiation techniques such as IMRT to reduce cardiac exposure, some degree of cardiac irradiation remains unavoidable in many lung cancer cases due to tumor proximity to mediastinal structures. Bitterman et al. demonstrated that a high percentage of patients with NSCLC, when switching to IMRT plans, were predicted to have reductions in MHD >4 Gy, at the expense of increasing low-dose lung exposure (^27^). However, CV events are still reported after thoracic RT using IMRT and VMAT, as illustrated in our study.

Tree-based classification models showed better performance than regression models in our study, as GBM outperformed all other models in AUC and RF had the highest sensitivity. These findings exactly align with Ladbury et al.’s results which demonstrated that GBM and RF outperformed LR models in predicting pulmonary, cardiac and esophageal toxicities in the secondary analysis of the Radiation Therapy Oncology Group (RTOG) 0617 trial (^28, 29^) using a cohort of 439 patients with NSCLC (^30^). Our ML models demonstrated limited ability to discriminate patients who would develop CVAE. Several factors could contribute to this modest performance. First, the small sample size limits the ability of complex models to capture subtle nonlinear relationships between predictors and outcomes. Second, CVAEs following CRT are multifactorial, and may depend on unmeasured cardiac substructure radiation doses, or biological, genetic, or treatment-related factors not included in the current dataset. Finally, heterogeneity of CV events grouped under the CVAE definition may reduce model specificity, as different pathophysiological mechanisms may underline ischemic, structural, and inflammatory cardiac complications. Niedzielski et al. demonstrated that regression models using whole heart and substructure dose had the highest performance (AUC=0.82) for prediction of ≥grade 2 post radiation pericardial effusion in a 141 cohort of patients with stage III NSCLC. Moreover, adding the substructure doses improved the performance of their predictive models (^31^).

Despite limited discrimination, feature importance analysis provided biologically plausible insights. Across both GBM and RF models, cardiac dose-volume metrics particularly heart V20, heart V40, heart V50, and MHD were consistently ranked among the most important predictors of CVAE. These findings support the well-established dose-response relationship between cardiac radiation exposure and CVAE risk (^9, 19, 32^). Previous studies in thoracic RT have demonstrated associations between higher heart dose metrics and increased risk of cardiac morbidity and mortality (^12, 33^).

Whole heart dosimetry has traditionally served as the primary method for quantifying cardiac risk following thoracic radiation. In a multivariate pooled analysis of LA-NSCLC patients undergoing definitive RT, Wang et al. reported two-year symptomatic cardiac event rates of 4%, 7%, and 21% for MHD of < 10 Gy, 10–20 Gy, and > 20 Gy, respectively, after accounting for baseline CV risk factors (^34^). Furthermore, in a long-term secondary analysis of RTOG 0617, Chun et al. demonstrated that increasing heart V20, V40, and V60 were all independently associated with worse OS (^35^). Most notably, heart V40 ≥ 20% vs < 20% showed median OS of 2.5 vs. 1.7 years, respectively (p < 0.001) (^35^). Current NCCN guidelines therefore recommend heart-dose volume constraints of V40 ≤ 20% and MHD ≤ 20 Gy for conventionally fractionated RT with concurrent chemotherapy (^36^). However, attention has increasingly turned toward cardiac substructure dosimetry, as whole heart metrics have proven to be inadequate surrogates for dose to specific structures, such as the left anterior descending artery (LAD) (^37^). Emerging evidence suggests that dose to certain substructures including the LAD, sinoatrial node, and cardiac chambers and valves may correlate with specific CVT such as ischemic heart diseases, arrhythmias, and CHF, respectively. These relationships are the focus of ongoing research and clinical trials aimed at establishing standardized organ-at-risk contouring guidelines and dose-volume constraints.

However, these associations should be interpreted cautiously as clinical and disease-related factors that may confound relationships between cardiac dosimetry and outcomes. Variables such as baseline CV risk burden, including composite indices like the MACE score, prior myocardial infarction, and smoking status, may independently influence both OS and CVAE. In addition, treatment-related factors, including exposure to systemic therapies with known cardiotoxic potential, and disease-specific factors such as tumor size and location, can affect both prognosis and the extent of cardiac irradiation.Systemic therapies themselves may contribute to CVT through biologic mechanisms independent of radiation dose. For example, increased expression of p16^^INK4a^^, a marker of cellular senescence and biologic aging, has been observed following cytotoxic chemotherapy and is associated with accelerated vascular aging and frailty (^38^). Similarly, cisplatin-based chemotherapy has been linked to endothelial dysfunction, vascular stiffness, and long-term atherosclerotic risk (^39^). Moreover, cisplatin-based regimens require aggressive intravenous hydration, which may exacerbate underlying cardiac dysfunction and precipitate clinical symptoms in patients with subclinical CVD as noticed in our clinics. Immunotherapy, particularly durvalumab consolidation following CRT has been associated with cardiac toxicities, including arrhythmias and new coronary artery diseases (^40^). Cardiac events may occur in 25% of patients with NSCLC received immunotherapy after CRT as demonstrated by Patel et al (^40^). Similarly, targeted therapies such as osimertinib, increasingly used in the adjuvant setting, are known to carry risks of CVT in EGFR-mutated NSCLC, particularly in patients with pre-existing comorbidities as our patients, necessitating routine cardiac monitoring (^41^). These treatment-related factors highlight the complex and multifactorial nature of CVT in lung cancer patients undergoing CRT. In parallel, surgical management of lung cancer, has also been associated with increased perioperative and longer-term cardiopulmonary complications and downstream mortality, particularly among patients undergoing more extensive resections or with significant baseline comorbidity burden (^42^). These overlapping determinants introduce the potential for residual confounding, complicating causal inference in dosimetric analyses and raising the possibility that heart dose may, in part, act as a surrogate for more aggressive disease in higher-risk patient populations rather than a direct mediator of outcomes.

Our results further suggest that even within a relatively small cohort, these dosimetric parameters remain key drivers of CVAEs. Age at diagnosis also emerged as an important predictor in both CVAE and mortality models. Older patients are more likely to have underlying CVD and reduced physiological reserve, making them more vulnerable to post-treatment complications. Tumor stage particularly Metastasis (M) stage was an important feature as well for CVAE prediction. Patients with metastasis have more advanced disease and receive chemotherapy as main therapy in the treatment plan. The combination of RT and chemotherapy increases the risk of severe cardiac events among patients with lung cancer as identified with Herbach et al. (^43^).

When we evaluated models using a reduced set of clinically and dosimetrically relevant variables, predictive performance remained similar to that observed with the larger feature set. This finding suggests that a limited number of variables, particularly age, smoking status, pre-existing CV comorbidity, and cardiac radiation dose metrics may capture much of the relevant predictive information. From a clinical perspective, this may support the development of simplified risk stratification tools that rely on readily available patient and treatment characteristics rather than large numbers of complex variables. Many prior studies in this field have been conducted in selected clinical trial populations, which often include healthier patients with fewer comorbidities and more controlled treatment conditions. In contrast, our study reflects a real-world Appalachian cohort characterized by a high burden of baseline CVD, smoking exposure, and socioeconomic disparities. This distinction is important, as real-world populations may experience higher rates of CVT, and different risk profiles compared to trial populations. Therefore, our findings may provide a more representative estimate of CV risk and enhance the generalizability of ML-based risk stratification in routine clinical practice.

In contrast to CVAE prediction, ML models demonstrated improved discrimination for mortality prediction, with RF achieving the highest AUC. Although these results still reflect modest predictive performance, they suggest that baseline clinical and dosimetric variables may contain meaningful information regarding OS indicating that mortality risk arises from the combined influence of demographic vulnerability, disease severity, and treatment-related cardiac exposure rather than from a single dominant factor. Cardiac dose volume metrics, particularly heart V50 (%), heart V20 (%), and heart V30 (%) were consistently ranked among the top contributors. These findings suggest that higher cardiac radiation exposure is associated with increased mortality risk in this cohort. These findings align with the results of Chan et al. which demonstrated that higher radiation dose to cardiac substructures are associated with decreased OS in early-stage NSCLC treated with SABR using neural networks (^44^). Speirs et al. also demonstrated that heart dose is associated with OS for patients with NSCLC treated with CRT (^32^). Additionally, Lee et al. demonstrated that explainable boosting machine (EBM), extreme gradient boosting and RF showed high performance to discriminate OS following RT using cardiac substructure radiation doses in lung cancers (^45^). Disease-related staging variables also contributed meaningfully indicating that tumor burden and metastatic spread influence mortality risk. Our results align with the findings of Ladbury et al., which identified group stage and N stage as the most important features for predicting ≥ grade 3 cardiac toxicity following RT in a secondary analysis of RTOG 0617 (^30^). Interestingly, baseline CV comorbidity and CVAE occurrence showed modest importance in mortality prediction, suggesting that baseline CVD and development of treatment-associated CVAEs are associated with increased mortality.

This study has several important clinical implications. First, our findings reinforce the importance of minimizing cardiac radiation exposure during thoracic RT whenever possible. Dose-volume parameters such as heart V20 and heart V50 are particularly relevant metrics for treatment planning and risk mitigation. Second, the integration of ML-based risk stratification tools could potentially assist clinicians in identifying high-risk patients who may benefit from closer cardiovascular monitoring or cardioprotective interventions during and after treatment. Lastly, the high prevalence of CVAEs observed in this Appalachian cohort highlights the need for multidisciplinary cardio-oncology approaches tailored to populations with elevated baseline CV risk.

## Limitations

Several limitations should be considered when interpreting these findings. The retrospective design and small sample size limit statistical power and generalizability. The study was conducted at a single institution, which may introduce institutional treatment patterns or referral biases. Additionally, CV events were aggregated into a single outcome variable, which may mask differences in mechanisms and predictors among distinct cardiac complications. Additionally, nearly all patients in this cohort received concurrent chemotherapy and a substantial proportion (77%) received immunotherapy, which may confound attribution of CVAE specifically to radiation exposure. Finally, external validation in independent cohorts was not performed and will be necessary before clinical implementation of these predictive models.

## Future directions

Future research should focus on externally validating these findings in larger, multi-institutional cohorts and incorporating additional biological and imaging biomarkers that may improve predictive performance. Integration of longitudinal cardiac imaging, cardiac substructure radiation dosimetrics, biomarkers of myocardial injury, and genomic susceptibility factors may help refine ML-based prediction of radiation-induced CVT. Furthermore, prospective studies evaluating ML-guided CV risk stratification in thoracic oncology could help determine whether these tools can meaningfully improve patient outcomes.

## 5 Conclusion

In conclusion, this study provides one of the first characterizations of cardiovascular adverse events after chemoradiation therapy and machine learning-based risk prediction in an Appalachian lung cancer population treated with definitive chemoradiotherapy. While machine learning models demonstrated modest predictive performance, cardiac radiation dose metrics and patient age consistently emerged as key predictors of both cardiovascular toxicity and mortality. These findings highlight the importance of cardiac dose optimization and continued development of predictive models incorporating multidimensional clinical, dosimetric, and imaging features to support personalized cardio-oncology care in high-risk populations.

## 6 Ethics approval and consent to participate

This retrospective study was approved by the Institutional Review Board (IRB) of West Virginia University (WVU): WVU Protocol # 2308839854. The study utilized existing clinical data extracted from electronic health records and given the retrospective nature of the study and the use of de-identified clinical data, the requirement for informed consent was waived.

## 7 Consent for publication

Not applicable as the study did not include identifiable individual patient data.

## 10 Authors’ contributions

Conceptualization: V.S., M.F.H and P.M.P.; methodology, data extraction, curation and data collection: V.S., J.A.S, T.N., J.C.K., A.N.J., M.T., R.A.S, J.R., C.M.B. and P.M.P; formal analysis, V.S., J.C.K. and J.A.S.; investigation, V.S., J.A.S., and J.C.K.; resources, R.A.S., D.A.C. and P.M.L.; writing original draft preparation, V.S. and J.A.S.; writing review and editing, J.R., R.A.S., C.M.B, R.R.R., D.A.C., M.F.H., and P.M.P.; visualization, V.S., J.A.S., and J.C.K.; supervision, M.F.H. and P.M.P.; funding acquisition, V.S., D.A.C., M.F.H. and P.M.P. All authors have read and agreed to the published version of the manuscript.

## Data Availability

The de-identified datasets and analysis code used in this study are publicly available in the Figshare repository at: https://doi.org/10.6084/m9.figshare.31900570 & DOI: 10.6084/m9.figshare.31901113. The shared dataset contains only de-identified information in accordance with institutional review board requirements.

## 11 Acknowledgements

We thank West Virginia University Cancer Institute for the Ignite Award and funds for supporting VS in her projects.

## 13 Use of Artificial Intelligence Statement

Artificial intelligence tools [ChatGPT (OpenAI, GPT-5.3 version)] were used to assist with language, grammar editing rephrasing, and code refinement during manuscript preparation. All analyses, interpretation of results, and final manuscript content were conducted, reviewed and verified by the authors, who take full responsibility for the accuracy and integrity of the work.

