## Supplemental File S1 for "Cardiovascular Adverse Events After Definitive Chemoradiotherapy for Lung Cancer in an Appalachian Population: Incidence and Machine Learning-Based Prediction"

| Feature | Importance_mean | Importance_std |
| --- | --- | --- |
| num__Heart V50 (%) | 0.054827 | 0.009584 |
| num__Age_at_Diagnosis | 0.035294 | 0.006659 |
| num__Heart V20 (%) | 0.028273 | 0.017337 |
| cat__M Stage | 0.027768 | 0.006962 |
| num__Mean Heart (Gy) | 0.023137 | 0.019315 |
| num__Heart V40 (%) | 0.022988 | 0.006574 |
| cat__Site of Primary | 0.016564 | 0.006655 |
| cat__N Stage | 0.011092 | 0.003925 |
| cat__Smoking | 0.008571 | 0.004572 |
| num__Heart V30 (%) | 0.00788 | 0.003152 |
| cat__T Stage | 0.006947 | 0.002981 |
| cat__Tumor_Histology | 0.00648 | 0.002239 |
| cat__Stage at Dx | 0.003978 | 0.002418 |
| cat__Sex | 0.002017 | 0.001291 |
| cat__Immunotherapy? | 0.000878 | 0.000761 |
| num__RT1 Dose (in Gy) | 0.000392 | 0.00165 |
| cat__Surgery? | 0.000317 | 0.000625 |
| cat__Race_Ethnicity | 0 | 0 |
| cat__Cardiovascular comorbidity | -0.0003 | 0.000967 |
| num__RT1 Fractions | -0.00131 | 0.001213 |

Supplementary Table 1: Feature importance of Random Forest model predicting Cardiovascular adverse events (CVAEs) in lung cancer after chemoradiation.

Supplementary Table 2: Feature importance of Gradient Boosting model predicting Cardiovascular adverse events (CVAEs) in lung cancer after chemoradiation.

| Feature | Importance_mean | Importance_std |
| --- | --- | --- |
| num__Heart V20 (%) | 0.140411 | 0.035114 |
| num__Age_at_Diagnosis | 0.028422 | 0.010304 |
| num__Heart V40 (%) | 0.010943 | 0.005886 |
| num__Mean Heart (Gy) | 0.007656 | 0.005362 |
| num__Heart V30 (%) | 0.003959 | 0.002893 |
| cat__Site of Primary | 0.001662 | 0.001471 |
| num__RT1 Fractions | 0 | 0 |
| num__RT1 Dose (in Gy) | 0 | 0 |
| cat__Sex | 0 | 0 |
| cat__Race_Ethnicity | 0 | 0 |
| cat__Smoking | 0 | 0 |
| num__Heart V50 (%) | 0 | 0 |
| cat__Cardiovascular comorbidity | 0 | 0 |
| cat__Tumor_Histology | 0 | 0 |
| cat__Stage at Dx | 0 | 0 |
| cat__T Stage | 0 | 0 |
| cat__N Stage | 0 | 0 |
| cat__M Stage | 0 | 0 |
| cat__Immunotherapy? | 0 | 0 |
| cat__Surgery? | 0 | 0 |

| Feature | Importance_mean | Importance_std |
| --- | --- | --- |
| num__Age_at_Diagnosis | 0.075735 | 0.026328 |
| num__Mean Heart (Gy) | 0.032805 | 0.008393 |
| num__Heart V50 (%) | 0.02802 | 0.005312 |
| num__Heart V20 (%) | 0.0256 | 0.009052 |
| num__Heart V30 (%) | 0.017851 | 0.00827 |
| cat__N Stage | 0.016222 | 0.005389 |
| num__Heart V40 (%) | 0.015588 | 0.005395 |
| cat__Cardiovascular comorbidity | 0.014864 | 0.008582 |
| cat__Site of Primary | 0.011968 | 0.003828 |
| cat__M Stage | 0.011923 | 0.004759 |
| cat__T Stage | 0.0094 | 0.003231 |
| cat__Tumor_Histology | 0.009186 | 0.002883 |
| cat__Stage at Dx | 0.00802 | 0.002759 |
| cat__CVT | 0.007523 | 0.004534 |
| num__RT1 Dose (in Gy) | 0.006391 | 0.003411 |
| cat__Smoking | 0.006312 | 0.001691 |
| cat__Sex | 0.006154 | 0.003376 |
| cat__Immunotherapy? | 0.004276 | 0.002437 |
| num__RT1 Fractions | 0.001731 | 0.000811 |
| cat__Race_Ethnicity | 0 | 0 |
| cat__Surgery? | -5.7E-05 | 0.00188 |

Supplementary Table 3: Feature importance of Random Forest model predicting mortality in lung cancer after chemoradiation.
